# Human cardiac stromal cells exposed to SARS-CoV-2 evolve into hyper-inflammatory/*pro*-fibrotic phenotype and produce infective viral particles depending on the levels of ACE2 receptor expression

**DOI:** 10.1101/2020.11.06.20226423

**Authors:** Alessandra Amendola, Gloria Garoffolo, Paola Songia, Silvia Ferrari, Giacomo Bernava, Paola Canzano, Veronika Myasoedova, Francesca Colavita, Concetta Castilletti, Giuseppe Sberna, Maria Rosaria Capobianchi, Marco Agrifoglio, Gualtiero I. Colombo, Paolo Poggio, Maurizio Pesce

## Abstract

Patients with severe respiratory syndrome caused by SARS-CoV-2 undergo cardiac complications due to hyper-inflammatory conditions. Although the presence of the virus has been detected in the myocardium of infected patients, and infection of cardiac cells may involve ACE2 receptor, the underlying molecular/cellular mechanisms are still uncharacterized. We analyzed expression of ACE2 receptor in primary human cardiac stromal cells using proteomic and transcriptomic methods before exposing them to SARS-CoV-2 *in vitro*. Using conventional and high sensitivity PCR methods, we measured virus production in the cellular supernatants and monitored the intracellular viral bioprocessing. We performed high-resolution imaging to show the sites of intracellular viral production. We finally used Q-RT-PCR assays to detect genes linked to innate immunity and fibrotic pathways coherently regulated in cells exposed to virus. Our findings indicate that human cardiac stromal cells have a susceptibility to SARS-CoV-2 infection and produce variable viral yields depending on the extent of cellular ACE2 receptor expression. Interestingly, these cells also evolved toward hyper-inflammatory/pro-fibrotic phenotypes independently of ACE2 levels, suggesting a dual cardiac damage mechanism that could account for the elevated numbers of cardiac complications in severe COVID-19 cases.

## Introduction

Since the beginning of the SARS-CoV-2 pandemic outbreak, a relatively high incidence of cardiac complications have been reported(Akhmerov and Marban, 2020; Libby, 2020). These range from elevation of cardiac damage markers such as circulating Troponin and BNP(Guo et al., 2020; Huang et al., 2020), to cardiac arrest(Baldi et al., 2020), cardiogenic shock(Tavazzi et al., 2020), myocarditis(Sala et al., 2020) and heart failure(Dong et al., 2020). The susceptibility of the myocardial tissue to SARS-CoV-2 infection(Escher et al., 2020; Tavazzi et al., 2020) has been inferred based on the expression of the Angiotensin-Converting Enzyme-2 (ACE2) receptor in various cardiac cell types(Zou et al., 2020), and the evidence that the virus interacts with this receptor *via* the Spike (S) protein, as a main cellular docking/internalization site(Wang, Q. et al., 2020). Although endothelial cells(Varga et al., 2020) and induced pluripotent cells (iPSCs)-derived cardiac myocytes(Sharma et al., 2020) are subject to SARS-CoV-2 infection, the relevance of the cardiac stroma in *pro*-inflammatory/pro-fibrotic evolution of the myocardium following acute injury(Humeres and Frangogiannis, 2019) suggests a potential involvement in myocardial acute inflammation and cardiac damage in COVID-19. This prompted us to investigate the effects of exposing primary cardiac derived stromal cells (cSt-Cs) to the virus *in vitro*.

## Results

### Variability in expression of ACE2 receptor in human cSt-Cs

Recently, individual variations in the level of *ACE2* mRNA expression have been reported in human myocardial cells, including cardiac fibroblasts, based on results of single cell RNA-sequencing experiments(Nicin et al., 2020). While this provides a rationale for the variable extent of cardiac damage observed in patients with COVID-19, it also suggests the involvement of cardiac stroma in the progression of the myocardial injury. Based on these evidences, we analyzed the levels of ACE2 protein expressed in 10 lines of human cardiospheres-derived cSt-Cs available to our laboratory (see Supplementary information for methodology of isolation and expansion)(Messina et al., 2004). **Table S1** and **Figure 1A** show, respectively, the characteristics of the cell donors, and the results of the ACE2 dosing in their cSt-Cs by Western Blotting and RT-qPCR. A relatively high variability in the expression of ACE2 protein was observed (**Figure 1A**), with no apparent relationship with demographic characteristics (e.g age), risk conditions (e.g. dyslipidemia, hypertension) or medication (e.g. anti-hypertensive treatment) (**Table S1**). On the other hand, protein levels were highly correlated with the levels of *ACE2* gene transcription, as verified by a linear regression analysis of the RNA/Protein expression data (**Figure 1A**). This indicates that the control of ACE2 expression in cST-Cs occurs at a transcriptional level.

**Figure 1.**
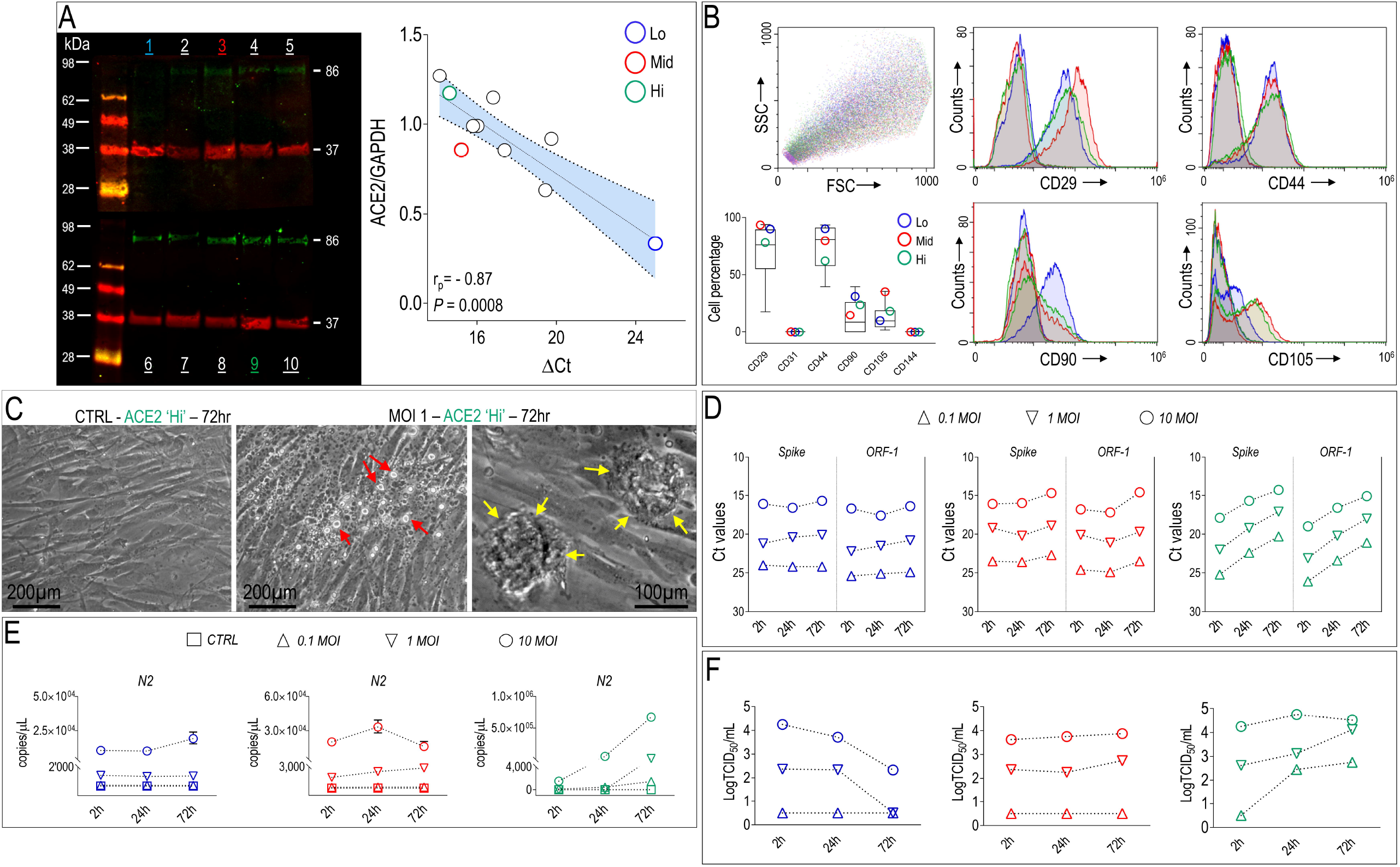
Expression of ACE2 and response of cST-Cs to infection with SARS-CoV-2. **(A)** Western analysis of ACE2 protein expression in 10 independent cST-Cs lines. ACE2 band is colored in green with a molecular weight (MW) ∼86 kDa. In red the GAPDH bands (MW 37 kDa) used for data normalization. On the right side, it is indicated the result of a linear regression analysis of protein/RNA data in the same cells, showing a high significance data correlation. In color it is indicated the 90% confidence interval. In both panels, numbers and symbols in color indicate, respectively, the data from the ‘Lo’ (blue), ‘Mid’ (red) and ‘Hi’ (green) cSt-Cs (see also **Table S1** for donors description). **(B)** Flow cytometry analysis of the cST-Cs with mesenchymal and endothelial markers. The FACS plots show the overlapped of the antigen expression profile of the cells from the three donors (each represented by their color code, see **Table S1**). The box-plot contains the min-max antigen expression data of the 10 cSt-Cs lines, with an indication of the three donors chosen for the experimental study represented by circles colored with the code adopted in **panel A** and **Table S1. (C)** Representative image of non-infected (CTRL, left panel) and SARS-CoV-2 infected ‘Hi’ cSt-Cs cells (1 MOI – 72 hours, center and right panels) showing vacuolization of the cytoplasm and release of microparticles (red arrows, center panel), and cytopathic effect characterized by cell rounding and wrinkling (yellow arrows, right panel). **(D)** Representation of the viral load in the cSt-Cs supernatant at various times post-infection with different MOI, as detected by conventional *Spike* and *ORF* RT-PCR amplification. Ct values were plotted on an inverse scale to appreciate the time dependent increase in viral concentration in supernatants of ACE2 ‘Hi’ and, at lower extent, of ‘Mid’ cells *vs*. the ‘Lo’ cells, which exhibited no variations compared to the level of the baseline at 2 hours. **(E)** dPCR detection of *N2* SARS-CoV-2 gene in cSt-Cs culture supernatant, indicative of absolute virus copy number expressed in copy number/µL. This analysis allowed to quantify with better resolution the release of viral particles by the three lines, and in particular, that even cells with the lowest level of ACE2 expression were able to produce viral particles in little amounts (see, e.g. the 72 hours - 10 MOI timepoint). **(F)** Titration of SARS-CoV-2 in the cSt-Cs supernatants was performed by VERO E6 cell line infection. In this experiment, the infectivity detected in the 2 hours supernatant (the experiment baseline) is indicative of the virus remnants in the medium after washing. As shown, the infectivity of the ‘Lo’ cells was decreasing over time, indicating absence of new virus production, while that of ACE2 ‘Hi’ and, at lower extent, of ACE2 ‘Mid’ cells, tended to increase, proving new viral production by cSt-Cs in the supernatant.

### ACE2 dependency of human cSt-Cs infection in vitro

Three cSt-Cs lines, indicated as ‘Hi’, ‘Mid’ and ‘Lo’ differing for the ACE2 protein expression level (**Figure 1A**), were chosen for the similarity of demographic (age, sex) and risk profile (diabetes, hypertension) of the donor subjects (**Table S1**). These cells were tested for expression of cardiac fibroblast/mesenchymal markers(Carlson et al., 2011). As shown in **Figure 1B**, expression of CD29 and CD44 was very similar, while a relatively higher variability was observed for expression of CD90 and CD105, typical markers of cardiac-resident mesenchymal cells(Smith et al., 2007). This variability, however, remained within the limits of the general variation in expression of mesenchymal markers in cells amplified from all donors (**Figure 1B**). Cells, finally, did not express endothelial markers CD31 and CD144 (**Figure 1B)**, excluding contamination by endothelial cells. To assess the tropism of SARS-Cov-2 for these cell lines, we exposed them to increasing amounts (multiplicity of infection – MOI, 0.1, 1, 10; see **Supplementary information** for details) of SARS-CoV-2 isolates(Capobianchi et al., 2020), and monitored the cytopathic effect and virological parameters from two to 72 hours. Exposure of the ACE2 ‘Hi’ cells to the virus induced a clear cytopathic effect that became visible already at two hours after the viral absorption, consisting of cell rounding and wrinkling, cytoplasmic volume reduction, and detachment. Vacuolization of the cells, and an increase in the number of secreted micro-vesicles was also noticed (**Figure 1C**). These effects were noticeable at lower extent, in ‘Mid’ cells, and undetectable in ACE2 ‘Lo’ cSt-Cs, even at 72 hours post-infection. In order to measure whether cells released viral particles in the supernatant, we performed conventional RT-PCR to detect *Spike* and *ORF* coding RNAs in the culture supernatants at 2, 24 and 72 hours post-infection (**Figure 1D**). Using this assay, ACE2 ‘Lo’ cells did not seem to release virus in the supernatant, as shown by the almost flat Ct curve as a function of time at all the considered virus concentrations. By contrast, ‘Mid’ and ‘Hi’ cells exhibited a clear increase of the Ct curve for both RNAs, evident for ‘Mid’ cells at 72 hours at 10 MOI, and for ‘Hi’ cells at 24 and 72 hours at all the MOIs, suggesting viral production. To confirm these data more quantitatively, we assessed the copy number of the virus in the supernatant using a digital-PCR (dPCR) amplification protocol using primers specific for the SARS-CoV-2 *N2* gene variant. Digital PCR methods, in our and others’ hands, are more sensitive than conventional PCR to detect viral copies in biological fluids with low viral titers ((Suo et al., 2020) and manuscript in preparation). As shown in **Figure 1E**, determination of the viral copy number was more precise with this technique. In particular, it was possible to appreciate that also the cell line expressing the lowest ACE2 levels produced viral particles in the supernatant (e.g. 72 hours, 10 MOI), even though their amount was almost three Log_10_ lower than those produced by the ‘Hi’ cells under the same conditions. To confirm that these viral particles are infective, we exposed the kidney-derived Vero E6 cell line (Keyaerts et al., 2005) to the supernatants of the infected cSt-Cs, followed by determination of the fifty-percent tissue culture infective dose (TCID_50_, see **Supplementary information**). Results of titration confirmed dPCR quantifications, showing a dose- and time-related increased infectivity above baseline recorded for ACE2 ‘Hi’ cellular supernatant and a gradual decrease of viral load in the supernatant of ACE2 ‘Lo’ cells (**Figures 1F, S1**). Taken together, these results suggest that cSt-Cs are susceptible to infection by SARS-CoV-2 in an ACE2-dependent manner and capable to support viral replication depending on the expression level of the receptor.

### Intracellular bioprocessing of SARS-CoV-2 in cSt-Cs

In order to investigate SARS-CoV-2 intracellular bioprocessing we first assessed the temporal dynamics of *E, N2* and *RdRP* viral transcripts in RNAs of cSt-Cs extracted cellular. In line with the previous results, clear differences were detected in the copy number of these genes in RNAs extracted from the different cell lines, with very limited number of copies/µL in ‘Lo’ cells at all the employed MOIs and MOI-dependent increases in ‘Mid’ and ‘Hi’ cells (**Figure 2A**). In order to find microscopic evidences of viral intracellular replication, we then performed immunofluorescence on ACE2 ‘Lo’ and ‘Hi’ infected cells using a human anti-SARS-CoV-2 serum from a COVID-19 convalescent patient (see **Supplementary information**), together with activated fibroblasts/myofibroblasts markers alpha-smooth muscle actin (αSMA) and/or Collagen-1 (Col1). Confocal microscopy imaging showed a very little proportion of ‘Lo’ cells exhibiting a SARS-CoV-2 labeling clearly above background fluorescence level observed in control cells (**Figure 2B**). By contrast, the number of ‘Hi’ cells labeled with the serum was markedly higher (**Figure 2C**). Infected cells exhibited a high number of intracellular viral particles (Figure **2D, E**) and extensive vacuolization of cSt-Cs cytoplasm, possibly indicating swelling of the endoplasmic reticulum (ER) associated to intense viral production (**Figure 2D**)(Romero-Brey and Bartenschlager, 2016) or the formation of a reticulo-vesicular ER networks supporting SARS-CoV-2 replication(Knoops et al., 2008). It was also interesting to observe that when cells exhibiting SARS-CoV-2 5 staining were found in contact with non-infected cells (**Figure 2F**), viral particles appeared to transit from the positive cell to the surrounding negative cells (see inset in **Figure 2F**). This suggests that SARS-CoV-2 may transfer from infected to uninfected cSt-Cs by direct cell- to-cell transfer, one of the modalities of viral intercellular propagation inside tissues(Zhong et al., 2013).

**Figure 2.**
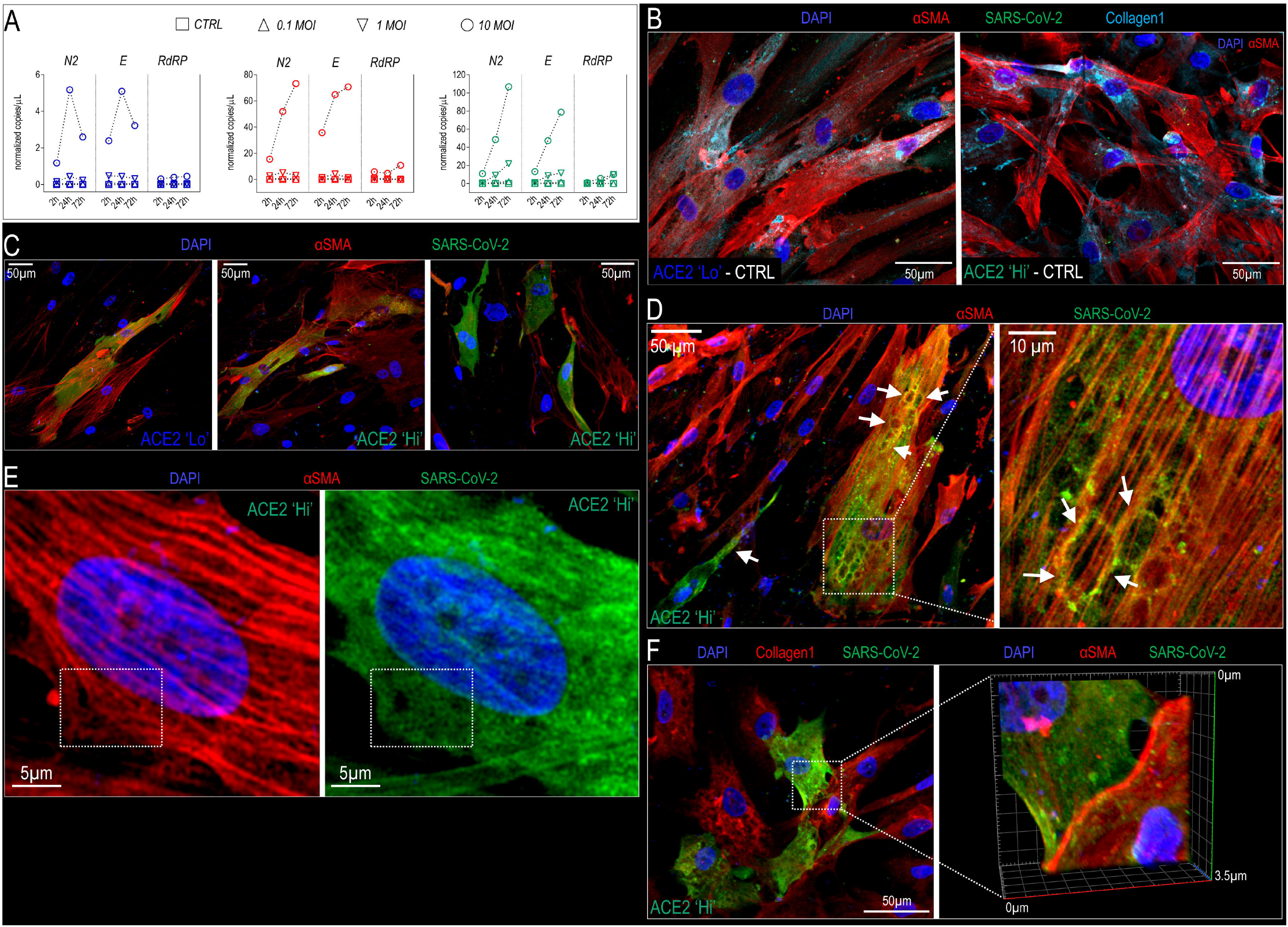
Intracellular bioprocessing of SARS-CoV-2 in cSt-Cs. **(A)** The intracellular bioprocessing of the virus was monitored by dPCR performed on cSt-Cs cellular RNAs extracted from cells at various times post-infection. In this case, the number of copies was normalized to *RpLP0* mRNA, used as a cellular internal control. Plots indicate the variation in *N2, E* and *RdRp* normalized copies/mL, indicative of the viral RNAs production inside cells, and provide an accurate estimate of the SARS-Cov-2 replication in cSt-Cs with different ACE2 expression (note the difference in the scale of the three plots in the panel). **(B)** Four-color immunofluorescence staining of uninfected ACE2 ‘lo’ and ‘Hi’ cells with nuclear staining (DAPI), αSMA, Collagen1 and SARS-CoV-2. In these cells, the expression of the markers was similar and the background color of SARS-CoV-2 staining was minimal. **(C)** Low-power magnification of triple-color stained cSt-Cs fixed 72 hours after exposure to SARS-CoV-2. ACE2 ‘Lo’ cells with a clear viral staining were very rare (left panel). The abundance of SARS-CoV-2^+^ cells was higher in ACE2’Hi’ cells; these cells exhibited a typical arrangement in small clusters among uninfected cells (mid and right panels). **(D)** Low- and high-power magnification of cells infected with SARS-CoV-2. In the left panel, note the presence of two cells with evident vacuolization of the cytoplasm (arrows). In the right panel, a high-power view of the zone encircled in the white square on the left. The oval structures surrounded by intense SARS-CoV-2 staining might be enlargements of the endoplasmic reticulum, one of the typical intracellular sites of virus assembly^22^. **(E)** High-power view of a cell showing αSMA^+^ stress fibers (left) and a cytoplasm densely packed with SARS-CoV-2 particles (insets). **(F)** In the panel on the left it is represented the low-power view of a cluster of cSt-Cs positive and negative for SARS-CoV-2 staining. Note in the panel on the right the 3D projection of a z-stacked acquisition of the region encircled in the white box in the left panel, where the boundary between the SARS-CoV-2^+^ and the SARS-CoV-2^-^ cells seems to be crossed by viral particles, indicative of possible direct cell-to-cell virus transmission.

### Pro-inflammatory and pro-fibrotic responses of cSt-Cs exposed to SARS-CoV-2

CSt-Cs have a central role in cardiac healing following acute injury, as they trigger the production of inflammatory cytokines and extracellular matrix remodeling enzymes necessary for recruitment of leukocytes and activation of the innate immunity process priming myocardium repair(Humeres and Frangogiannis, 2019). Since SARS-CoV-2 infection causes sharp upregulation of inflammatory cytokines in target organs through infection-dependent and innate immunity signaling mediated by Toll-like receptors(Sallenave and Guillot, 2020), it is possible that cardiac inflammation observed in COVID-19 patients results from a combination of the systemic ‘cytokine storm’ and a direct inflammatory response by cardiac-resident cells(Guzik et al., 2020). In order to assess this hypothesis, we tested the effects of the virus on activation of inflammatory factors and genes potentially involved in cardiac fibrosis^16^. We therefore analyzed the expression of genes involved in innate immune response and cardiotoxicity using RNAs extracted from the three cell types infected with SARS-CoV- 2 for 2, 24 and 72 hours. As shown in **Figure 3A**, unsupervised clusterization of the differentially expressed genes (**Table S4**) revealed a time-dependent regulation trend, indicative of convergence of the three cell lines toward a similar response to viral exposure, independent of ACE2 expression level. The genes that were significantly upregulated at 2 hours post infection indicate an early response of cSt-Cs to stress conditions determined by exposure to the virus. For example, *HSPH1 (hsp110)* is a heat shock protein strongly upregulated in response to coronaviruses exposure and in particular to their Envelope (E) proteins(DeDiego et al., 2011), while *FOSL1* is a transcription/cellular factor engaged in Interferon signaling in responses to viral infection(Cai et al., 2017). It was remarkable to note that some of the transcripts significantly upregulated at 72 hours in response to virus encode for, ***i)*** a membrane adhesion protein involved in cell-to-cell intercellular viral transmission (e.g. ICAM1) (Bracq et al., 2018); ***ii)*** a chemokine with potent *pro*-inflammatory effects (CCL7/MCP3) in COVID-19 (Vaninov, 2020; Yang, Y. et al., 2020); and ***iii)*** transcriptional regulators EGR1 and STAT1 involved, respectively, in SARS-CoV-related TGF-β1 signaling (Li et al., 2016) and immune response in COVID-19 patients (Zhu et al., 2020). We finally investigated regulation of mRNAs encoding for key factors involved in COVID-19 ‘cytokine storm’ and cardiac inflammatory/fibrotic responses(Adao and Guzik, 2020; Akhmerov and Marban, 2020; Ammirati and Wang, 2020). Results of RT-qPCR clearly indicated that cells from the three cell lines responded similarly to viral exposure with a time-dependent upregulation of pro-fibrotic genes *CTGF, ACTA2, Col1A* and *Col3A* and of inflammatory cytokines *IL-1*β and *CCL2* (*MCP1*) and, to a lower extent, *IL-6* mRNAs irrespective of ACE2 expression levels (**Figure 2B, C**). Together, these results highlight an additional cardiac pathogenesis mechanism by SARS-CoV-2 independent of ACE2 expression, consisting of substantial upregulation of genes involved in response to viral infection, intercellular virus transmission and related to innate immunity signaling and fibrotic activation.

**Figure 3.**
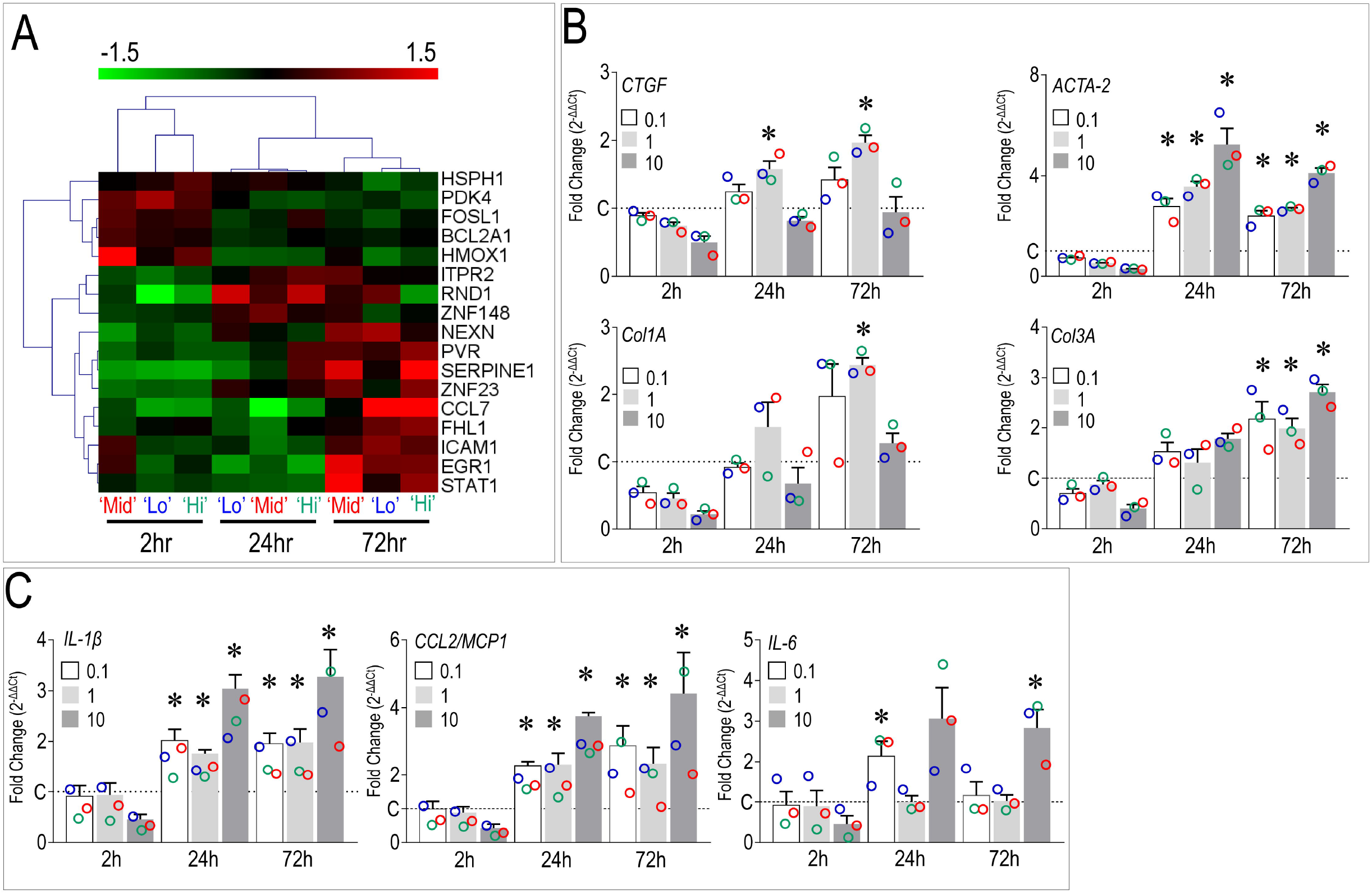
Innate immnunity and pro-fibrotic responses in cSt-Cs exposed to SARS-CoV-2. **(A)** Two RT-qPCR gene arrays containing primers for amplification of 164 transcripts potentially involved in cardiotoxicity and innate immune signaling were employed to assess changes in gene expression consequent to exposure of the three cSt-Cs lines to SARS-CoV-2 (10 MOI). After identification of the genes significantly changed in their expression levels at 2, 24 and 72 hours post-infection with a *P*-value < 0.05 by ANOVA, an unsupervised data clusterization was conducted. Results showed a coordinated clusterization of transcripts of the three cell lines, with groups of genes up/downmodulated at different time points. Raw data of this experiment are represented in **Table S4. (B, C)** The expression of genes typically involved in cSt-Cs pro-fibrotic activation (*CTGF, ACTA2, Col1A and Col3A*) and the SARS-CoV-2 cytokine storm (*Il-*β, *CCL2/MCP1, IL-6*) was finally investigated by single RT-qPCR analyses on cellular RNAs at the three time points and at all the tested viral concentrations. As shown, the expression of the pro-inflammatory cytokines was consistently upregulated in the three lines above the level of uninfected cells already at 24 hours of expression. Again, upregulation of transcription of these genes appeared independent of the level of ACE2 expression, as evidenced by overlapping the values of the gene expression fold changes in the individual cells (color coded as in **Fig 1A** and **Table S1**) to the bar graphs indicating the average and the standard error of the data. In both panels * indicate *P* < 0.05 statistical significance by one way ANOVA analysis (repeated measures) of the ΔCt values for each gene at all the viral concentrations used for the infection *vs*. uninfected cells at 24 and 72 hours post infection.

## Discussion

Experimental evidences have shown the susceptibility of cells expressing ACE2 receptor to direct infection by SARS-CoV-2 with implications for the multi-organ disease characterizing the COVID-19 pandemic outbreak. This includes endothelial(Varga et al., 2020), kidney and urogenital tract cells(Wang, S. et al., 2020), enterocytes(Lamers et al., 2020), and a variety of human iPSCs-derived cell types(Yang, L. et al., 2020), including cardiac myocytes. As of today, despite the numerous reports showing extensive cardiac damages consequent to infection and the presence of the virus in myocardial biopsies(Tavazzi et al., 2020; Wenzel et al., 2020), there is still uncertainty about the underlying mechanisms(Guzik et al., 2020). As outlined in various cardiology-oriented reviews on COVID-19 pathophysiology(Akhmerov and Marban, 2020; Libby, 2020), the heart could be affected by cumulative effects of the cytokine storm elicited by innate immunity activation(Sallenave and Guillot, 2020), as well as of *in situ* cytopathic effect determined by direct infection and replication of the virus in the myocardium(Tavazzi et al., 2020; Wenzel et al., 2020). As discussed, these effects may cause a post-pandemic increase of heart failure (Thum, 2020). In the present study, we provide the first proof-of-concept that cells of the human myocardial stroma are susceptible to infection and permissive for intracellular replication of SARS-CoV-2. We also show that virus infectivity and productivity depend on the level of the ACE2 receptor, thus confirming the relevance of individual variations in the expression of this receptor for potential responses to infection (Nicin et al., 2020). Interestingly, exposure of the cells to SARS-CoV-2 was also able to elicit innate immunity and *pro*-fibrotic responses in cSt-Cs. This effect was not related to ACE2 expression, but to infection-independent mechanisms. In conclusion, based on the data of the present report, we hypothesize that SARS-CoV-2 causes cardiac complications with potentially cumulative effects of ***i)*** an intra-myocardial cytopathic effects due to viral replication in the stromal component connected to 7 individual ACE2 protein levels and, ***ii)*** an ACE2-independent innate immunity response boosting myocardial inflammation and fibrosis(Ammirati and Wang, 2020).

## Data Availability

Data will made available at a public data repository upon request after publication.

## Acknowledgements

This work has been financed by a Grant from Regione Lombardia (POR FESR 2014-2020-LINEA 2A COVID-grant no. 1850333) granted to M.P. and A.A. M.P. and A.A. are supported by Institutional grants (Ricerca Corrente, 5 per 1000) issued by Italian Ministry of Health. A.A. was supported also by COVID-2020-12371817 and COVID-2020-12371675 program grants from Italian Ministry of Health.

## Notes

### Competing Interest Statement

The authors have declared no competing interest.

### Clinical Trial

N/A

### Author Declarations

Derivation of cells from human atrial appendage was approved by the Ethical Committee at Centro Cardiologico Monzino, IRCCS, Milano. A written agreement was obtained by each patient before collection of the material, during routine cardiac surgery interventions.

