## supplementary information for "Human cardiac stromal cells exposed to SARS-CoV-2 evolve into hyper-inflammatory/*pro*-fibrotic phenotype and produce infective viral particles depending on the levels of ACE2 receptor expression"

### Ethical Information

Derivation of cells from human atrial appendage was approved by the Ethical Committee at Centro Cardiologico Monzino, IRCCS; Milan. Each patient, admitted for routine cardiac surgery interventions signed an informed consent to authorize collection of the material.

**Table S1.** Patients demographic and risk characteristics, with an indication of the anti-hypertensive therapy. Data in the columns on the right indicate the normalized ACE2/GAPDH protein and the ACE2 mRNA (ΔCt values) expression levels, as detected by western analysis and RT-qPCR, respectively. In blue, red and green colors, the subjects that were chosen as ACE2 'Lo', 'Mid' and 'Hi' cST-Cs donors, respectively.

| Patient | Sex | Age range | Diabetes | Dyslipidemia | Hypertension | Therapy | ACE2 normalized level (WB) | ACE2 mRNA (ΔCt) |
|---|---|---|---|---|---|---|---|---|
| **1** | **M** | 70-80 | **Y** | **Y** | **Y** | **Perindopril** | **0.334** | **24,984** |
| 2 | M | 70-80 | N | Y | Y | Enalapril | 0.989 | 16.022 |
| **3** | **M** | 70-80 | **Y** | **N** | **Y** | **-** | **0.854** | **15.215** |
| 4 | M | 70-80 | N | Y | Y | Losartan | 0.989 | 15.794 |
| 5 | M | 70-80 | Y | N | Y | Olmesartan | 1.270 | 14.113 |
| 6 | M | 70-80 | N | N | Y | - | 0.854 | 17.388 |
| 7 | M | 60-70 | N | Y | Y | Perindopril | 0.632 | 19.443 |
| 8 | M | 50-60 | Y | Y | Y | Ramipril | 1.147 | 16.815 |
| **9** | **M** | 70-80 | **Y** | **Y** | **Y** | **Losartan** | **1.173** | **14.607** |
| 10 | F | 50-60 | N | N | Y | Perindopril | 0.918 | 19.749 |

### Preparation of cSt-Cs and antigenic/proteomic characterization

For preparation of human cardiac stromal cells, human right atrial appendage biopsies, obtained from patients undergoing cardiac surgery, were subjected to mechanical fragmentation and enzymatic digestion, and cultured for 4 weeks in Iscove's modified Dulbecco's medium (IMDM) supplemented with 20% Hyclone characterized Fetal Bovine Serum (FBS), 1% penicillin-streptomycin, 1% L-glutamine and 0.1mM2-mercaptoethanol. Explant-derived cells were plated on poly-D-lysine coated wells in the following media to obtain primary cardiospheres: 35% IMDM/65% DMEM/F-12 Mix, 3.5% Hyclone characterized FBS, 1% penicillin-streptomycin, 1% L-glutamine, 0.1 mM 2-mercaptoethanol, 1 unit/ml thrombin, 1:50 B-27, 80 ng/ml bFGF, 25 ng/ml EGF, and 4 ng/ml cardiotrophin-1 (all Peprotech). cSt-Cs were then derived by dissociating cardiospheres followed by plating onto Fibronectin-coated dishes. For antigenic characterization, cST-Cs suspensions (passages 1-4) were incubated with a combination of fluorochrome-conjugated antibodies against CD29, CD31, CD44, CD90, CD105 and CD144 (or the corresponding Isotype antibodies; all from BD Biosciences) for 15 minutes at room temperature (RT). Cells were analyzed with Gallios Flow Cytometer (Beckman Coulter) equipped with Kaluza analysis software (Beckman Coulter).

### Western blot

Cells were lysed in a buffer containing 10 mM Tris-Cl, pH 7.4, 150 mM NaCl, 5 mM EDTA, 1% (v/v) Triton X-100, 1% (w/v) sodium deoxycholate, 0.1% (w/v), sodium dodecyl sulfate and 1% (v/v), protease and phosphatase inhibitor mixture (Sigma-Aldrich). Whole cell lysates were sonicated, centrifuged for 15 min at14 000 g; cell supernatants were then collected. Proteins were quantified by BCA protein assay kit (Pierce Chemical Co). Cell lysates (30 μg per lane) were diluted in Laemli sample buffer, heated at 95 °C for 5 minutes, run onto 4-12% gradient SDS polyacrylamide gels (Invitrogen), and transferred to nitrocellulose membranes. The blots were blocked with Tris Buffered-saline containing 5% (w/v) BSA at RT for 1 hour. Overnight incubation at 4 °C with ACE2 and GAPDH primary antibodies (Novus Biologicals; Santa Cruz Biotechnology) was performed. Membranes were incubated with appropriate secondary antibodies for 20 minutes. Images were acquired with LI-COR Odyssey and band intensities were quantified using ImageJ software.

**Preparation of SARS-CoV-2 isolates**

Viral stock used to infect cSt-Cs was the first Italian SARS-Cov-2 isolate (named 2019-nCoV/Italy-INMI1 and available from EVAg, Ref-SKU: 008V-03893) obtained from a patient hospitalized at National Institute for Infectious Diseases "Lazzaro Spallanzani" (INMI) in Rome. Virus was expanded on Vero E6 cells (ATCC) maintained in Modified Eagle Medium (MEM) supplemented with 10% heat inactivated FBS, 2 mM L-glutamine, and 1% penicillin/streptomycin solution at 37°C, in a humidified atmosphere of 5% CO2. Virus titer was determined in culture supernatants as fifty-percent tissue culture infective dose (TCID50/mL), according to Reed and Muench method[16].

**SARS-CoV-2 infection of cSt-Cs**

For infection with SARS-CoV-2, the cSt-Cs lines were seeded in 6-well culture plates. After reaching cell confluence, they were exposed to Vero E6 supernatant containing SARS-CoV-2 at the multiplicity of infection (MOI) of 0.1, 1 and 10 for 2 hour at 37°C. Supernatant was then removed and, after three washes with DPBS, fresh culture medium was added for 3 days. cSt-Cs supernatants were collected at 2, 24 and 72h post infection to assess the kinetics of viral replication. Supernatants were in part lysed in TRIZOL for molecular detection of the virus by RT-qPCR and digital PCR immediately and in part frozen at -80 °C for further investigation. At the time of supernatant collection, cells were lysed in TRIzol (Invitrogen) to recover cellular RNAs and perform the analyses on cellular and viral RNAs (see below).

**Titration of SARS-CoV-2 produced by the cST-Cs in Vero E6 cells**

The infectivity of the virus produced by cSt-Cs collected at 2h, 24h and 72h post infection was measured by Reed and Muench method on Vero E6 cells [16]. Vero E6 cells were seeded in 96-wells culture plates. Ten fold dilutions of cellular supernatants were prepared in culture medium supplemented with 2% heat-inactivated FBS and 0,05ml of each dilution was added to the cells. The plates were incubated for 48-72h at 37 °C. The kinetics of virus infecgtion was followed by PFU for 72h and expressed as fifty-percent tissue culture infective dose (TCID50/mL).

**RT-qPCR in cellular lysates**

Total RNA from infected cells at various times with different MOI was extracted using TRIzol (Invitrogen), quantified with NanoDrop-1000 spectrophotometer (Thermo Fisher Scientific). Superscript III (Thermo Fisher Scientific) was used for reverse transcription. Quantitative real-time PCR analysis were performed with Power SYBR Green PCR Master Mix (Applied Biosystems) in an ABI 7900 Fast thermal cycler to detect *ACE2*, inflammatory cytokines and *pro*-fibrotic gene amplification products (primers details in **Table S2**). The reported expression levels were calculated relative to GAPDH mRNA, used as an internal standard control. The fold change of the genes in infected cells *vs.* control samples was calculated as $2^{-\Delta\Delta CT}$ and the statistical analysis was done on the ΔCT values.

.

**Table S2.** List of primers used to perform RT-qPCR analyses on cellular extracts.

| Gene | Sequence |
|---|---|
| *hACE2* | Forward: 5'- GCCACTGCTCAACTACTTTG -3'<br>Reverse: 5'- GCTTATCCTCACTTTGATGCTTTG -3' |
| *hIL-1β* | Forward: 5'- CAGCCAATCTTCATTGCTCAAG -3'<br>Reverse: 5'- GAACAAGTCATCCTCATTGCC -3' |

| | |
|---|---|
| *hIL-6* | Forward: 5'- GCAGATGAGTACAAAAGTCCTGA -3'<br>Reverse: 5'- TTCTGTGCCTGCAGCTTC -3' |
| *hCCL2* | Forward: 5'- AGCAGCCACCTTCATTCC -3'<br>Reverse: 5'- GCCTCTGCACTGAGATCTTC -3' |
| *hACTA2* | Forward: 5'- AGAGTTACGAGTTGCCTGATG -3'<br>Reverse: 5'- CTGTTGTAGGTGGTTTCATGGA -3' |
| *hCTGF* | Forward: 5'- AGGAGTGGGTGTGTGACGA -3'<br>Reverse: 5'- CCAGGCAGTTGGCTCTAATC -3' |
| *hCOL1A1* | Forward: 5'- GGACACAGAGGTTTCAGTGG -3'<br>Reverse: 5'- CCAGTAGCACCATCATTTCC -3' |
| *hCOL3A* | Forward: 5'- AGCTACGGCAATCCTGAACT -3'<br>Reverse: 5'- GGGCCTTCTTTACATTTCCA-3' |
| *hGAPDH* | Forward: 5'- AATCCCATCACCATCTTCCAG -3'<br>Reverse: 5'- AAATGAGCCCCAGCCTTC -3' |

**mRNA analysis with gene arrays**

Human Cardiotoxicity RT2 Profiler PCR Array (PAHS-095ZA, Qiagen) and Human NF-κB Signalling pathway plus RT2 Profiler PCR Array (PAHS-025YE, Qiagen) were performed on an Applied Biosystems (ABI) 7900 according to the manufacturer's instructions using 400 ng of each RNA preparation. Data were expressed as fold changes compared to the control condition, calculated with the 2-ΔΔCt method (i.e., using the cells harvested before infection as internal calibrator). Differential and clustering analyses were performed using algorithms incorporated in the open-source software MultiExperiment Viewer (MeV) version 4.9. Differences in gene expression were identified by one-way ANOVA: P-values computed from the F-distribution <0.05 were deemed as significant. Hierarchical cluster analysis was performed to visualize the similarities and differences in gene expression among the time points. Normalized gene expression values were mean-centered, scaled and clustered by Pearson's correlation and average linkage.

**Quantification of SARS-CoV-2 in cST-Cs supernatants and cell extracts by conventional and digital PCR methods**

For an initial evaluation of the virus released by infected cST-Cs in culture supernatant during the time course (2, 24 and 72h p.i.) we used a diagnostic RT-qPCR assay (Simplexa™ COVID-19 Direct assay, Diasorin S.p.A) targeting *Spike* (S) and *ORF1ab* SARS-CoV-2 coding sequences. Data were expressed as Ct values of each gene as detected in the supernatant of the cells at each time point.

For viral RNA quantification in cellular extracts and supernatants, a one-step chip-based digital PCR was performed on a QuantStudio 3D Digital PCR System platform composed by the QuantStudio 3D Instrument, the Dual Flat Block GeneAmp PCR System 9700 and the QuantStudio 3D Digital PCR Chip Loader (Thermo Fisher Scientific). TaqMan Fast Virus 1-Step Master Mix (Applied Biosystems) was used in combination with TaqMan primers and FAM probe for viral RNA (**Supplementary Table S3**), purchased from Integrated DNA Technologies.

Regarding the supernatant viral RNA quantification, 5 µL of the extracted extracellular RNA were analyzed with primes and probe for viral N gene. Concerning the intracellular viral RNA quantification, 20 ng of extracted cellular RNA were analyzed with primers and probes for viral N, E, and RdRp genes. Human RNAse P gene was used as internal amplification control. Reverse transcription was performed at 50 °C for 10 minutes, followed by inactivation/initial denaturation at 96 °C for 5 minutes. PCR cycling consisted of a denaturation step at 98 °C for 30 seconds followed by annealing/extension at 56 °C for 1 minute (40 cycles) and a final extension at 60 °C for 5 minutes. Analyses were executed with the online version of the QuantStudio 3D

AnalysisSuite (Thermo Fisher Scientific Cloud) following the Manufacturer's instructions.

**Table S3.** List of primers and probes used for the viral RNA evaluation with dPCR.

| Gene | Type | Concentration | Number |
|---|---|---|---|
| nCOV_N | Forward Primer | 50 nmol | 10006824 |
| nCOV_N | Reverse Primer | 50 nmol | 10006825 |
| nCOV_N | Probe | 25 nmol | 10006826 |
| E_Sarbeco | Forward Primer | 50 nmol | 10006888 |
| E_Sarbeco | Reverse Primer | 50 nmol | 10006890 |
| E_Sarbeco | Probe | 25 nmol | 10006892 |
| RdRP_SARSr | Forward Primer | 50 nmol | 10006860 |
| RdRP_SARSr | Reverse Primer | 50 nmol | 10006881 |
| RdRP_SARSr | Probe | 25 nmol | 10006886 |
| RNase P | Forward Primer | 50 nmol | 10006827 |
| RNase P | Reverse Primer | 50 nmol | 10006828 |
| RNase P | Probe | 25 nmol | 10006829 |

**Immunofluorescence**

After fixation with 4% paraformaldehyde, cells were permeabilized for 1 hour at RT with PBS containing 3% (w/v) BSA and 1% (v/v) Triton X-100, followed by incubation with COVID-19 patient derived-serum and the primary antibodies against αSMA (DAKO) and Col1 (Rockland) at 4°C overnight. Cells were then incubated for 1 hour at RT with the appropriate secondary antibodies, Phalloidin-TRITC and DAPI nuclear dye. Digital images were obtained using an LSM710 confocal scanning microscope (Carl Zeiss) equipped with the AiryScan super resolution system.

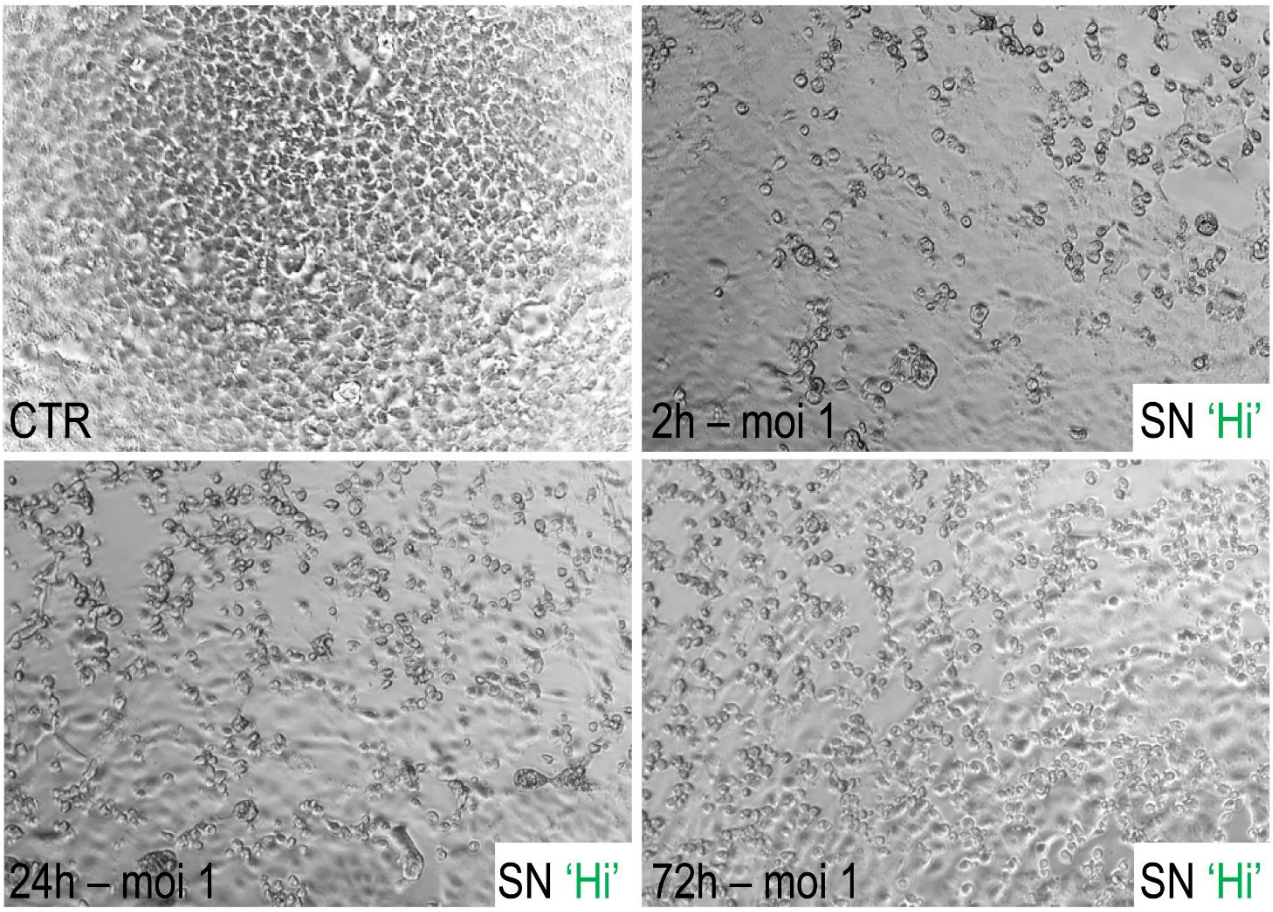


**Figure S1.** VERO E6 cells were exposed to culture supernatants of cSt-Cs infected with SARS-CoV-2 to perform viral titration. The picture shows the cytopathic effect induced by SARS-CoV-2 contained in the ACE2 'Hi' cells supernatant after infection (1 MOI) for 72 hours.

**Table S4.** List of the transcripts significantly affected in cSt-Cs in response to SARS-CoV-2 exposure. Data are expressed as fold change of each transcript in the given cell/infection condition/time vs. the levels of the same gene expressed in uninfected cells. The statistical significance (P-value) of the data has been calculated with **ANOVA.**

| Gene | Lo 2h | Mid 2h | Hi 2h | Lo 24h | Mid 24h | Hi 24h | Lo 72h | Mid 72h | Hi 72h | Mean 2h | Mean 24h | Mean 72h | *P*-value |
|---|---|---|---|---|---|---|---|---|---|---|---|---|---|
| PDK4 | 1,56 | 1,16 | 1,06 | 0,61 | 0,19 | 0,13 | 0,40 | 0,27 | 0,13 | 1.26 ± 0.26 | 0.31 ± 0.26 | 0.27 ± 0.14 | 0,0029 |
| EGR1 | 0,52 | 1,45 | 0,92 | 0,21 | 0,60 | 0,12 | 1,77 | 2,46 | 1,80 | 0.96 ± 0.47 | 0.31 ± 0.26 | 2.01 ± 0.39 | 0,0045 |
| STAT1 | 0,79 | 1,08 | 0,94 | 0,78 | 0,73 | 0,66 | 1,44 | 2,72 | 2,09 | 0.94 ± 0.15 | 0.72 ± 0.06 | 2.08 ± 0.64 | 0,0097 |
| NEXN | 1,20 | 0,70 | 1,01 | 1,80 | 1,46 | 1,22 | 2,52 | 2,32 | 1,75 | 0.97 ± 0.25 | 1.49 ± 0.29 | 2.20 ± 0.40 | 0,0097 |
| BCL2A1 | 0,62 | 0,84 | 0,54 | 0,11 | 0,16 | 0,40 | 0,22 | 0,29 | 0,37 | 0.67 ± 0.16 | 0.22 ± 0.16 | 0.29 ± 0.08 | 0,0138 |
| ZNF23 | 1,06 | 0,95 | 0,99 | 1,88 | 1,59 | 1,85 | 1,45 | 2,24 | 2,37 | 1.00 ± 0.06 | 1.77 ± 0.16 | 2.02 ± 0.50 | 0,0148 |
| ITPR2 | 0,76 | 1,02 | 1,05 | 1,39 | 1,76 | 2,04 | 1,54 | 2,01 | 1,50 | 0.94 ± 0.16 | 1.73 ± 0.33 | 1.68 ± 0.28 | 0,0189 |
| ICAM1 | 0,87 | 1,62 | 0,82 | 1,01 | 0,52 | 0,78 | 2,00 | 1,60 | 1,74 | 1.10 ± 0.45 | 0.77 ± 0.25 | 1.78 ± 0.20 | 0,0209 |
| SERPINE1 | 0,78 | 0,88 | 0,74 | 1,01 | 1,46 | 2,28 | 1,89 | 3,08 | 3,78 | 0.80 ± 0.07 | 1.58 ± 0.64 | 2.92 ± 0.96 | 0,0218 |
| HSPH1 | 1,12 | 0,95 | 1,42 | 1,03 | 1,14 | 0,97 | 0,23 | 0,77 | 0,59 | 1.16 ± 0.24 | 1.05 ± 0.09 | 0.53 ± 0.27 | 0,0246 |
| CCL7 | 0,99 | 1,54 | 1,05 | 1,47 | 0,46 | 1,16 | 4,15 | 1,91 | 4,78 | 1.19 ± 0.30 | 1.03 ± 0.52 | 3.61 ± 1.51 | 0,0258 |
| PVR | 1,25 | 0,94 | 1,03 | 1,03 | 1,35 | 2,01 | 1,85 | 2,03 | 2,35 | 1.07 ± 0.16 | 1.46 ± 0.50 | 2.08 ± 0.25 | 0,0288 |
| RND1 | 0,55 | 1,72 | 1,12 | 3,27 | 2,45 | 3,15 | 2,65 | 2,35 | 1,18 | 1.13 ± 0.59 | 2.96 ± 0.44 | 2.06 ± 0.78 | 0,0307 |
| FHL1 | 1,06 | 0,74 | 1,21 | 0,65 | 0,40 | 1,13 | 1,72 | 1,26 | 1,91 | 1.00 ± 0.24 | 0.73 ± 0.37 | 1.63 ± 0.33 | 0,0338 |
| ZNF148 | 1,03 | 0,96 | 1,12 | 1,66 | 1,99 | 1,51 | 0,93 | 1,60 | 1,31 | 1.04 ± 0.08 | 1.72 ± 0.25 | 1.28 ± 0.34 | 0,0369 |
| FOSL1 | 0,91 | 1,20 | 1,01 | 0,30 | 0,49 | 0,95 | 0,20 | 0,45 | 0,55 | 1.04 ± 0.15 | 0.58 ± 0.33 | 0.40 ± 0.18 | 0,0382 |
| HMOX1 | 0,95 | 2,64 | 1,41 | 0,29 | 0,31 | 0,35 | 0,14 | 0,93 | 0,48 | 1.67 ± 0.87 | 0.32 ± 0.03 | 0.52 ± 0.40 | 0,0492 |
